# Differences in Plasma Metabolites Related to Alzheimer’s Disease, APOE-ε4 status and Ethnicity

**DOI:** 10.1101/2020.01.10.20016741

**Authors:** Badri Vardarajan, Vrinda Kalia, Jennifer Manly, Adam Brickman, Dolly Reyes-Dumeyer, Rafael Lantigua, Iuliana Ionita-Laza, Dean P. Jones, Gary W. Miller, Richard Mayeux

## Abstract

**INTRODUCTION:** We investigated metabolites in plasma to capture systemic biochemical changes associated with Alzheimer’s disease (AD).

**METHODS:** Metabolites in plasma were measured in 59 AD cases and 60 healthy participants of African American (AA), Caribbean Hispanic (CH) and non-Hispanic white (NHW) ancestry using untargeted liquid-chromatography based ultra-high resolution mass spectrometry. Metabolite differences between AD and healthy, ethnic groups and APOEε4 status were analyzed. Untargeted network analysis identified pathways enriched in AD-associated metabolites.

**RESULTS:** 5,929 annotated metabolites were measured. PLS-DA analysis inferred that AD clustered separately from healthy controls (AUC=0.9816); discriminating pathways included glycerophospholipid, sphingolipid and non-essential amino acid (alanine, aspartate, glutamate) metabolism. Metabolic features in AA clustered differently from CH and NHW (AUC=0.9275), and differed between *APOE*ε4-carriers and non-carriers (AUC=0.9972).

**DISCUSSION:** Metabolites, specifically lipids, were associated with AD, *APOE*ε4 and ethnic group. Metabolite profiling could identify perturbed AD pathways, but genetic and ancestral background need to be considered.

## Introduction

The understanding of the pathogenesis of Alzheimer’s disease (AD) is incomplete. Genetic and molecular mechanisms have been proposed, but no single gene variant or molecular mechanism can fully explain the complexity of this disorder. Thus, it is likely that genetic variants affect downstream metabolic pathways. Finding systemic molecular changes between disease and healthy states could identify biological mechanisms, potentially leading to early diagnosis and therapeutic development and possibly successful interventions.

Metabolomics is a method of deep molecular phenotyping that represents the underlying biochemical and physiological layer of the genome, transcriptome and proteome. Metabolomics provides a precise and comprehensive analysis of phenotypic abnormalities in which the individual components are physiologically described. This snap shot of the metabolic state of individuals is possible due to the advances in high-resolution mass spectrometric platforms with high sensitivity and specificity^1^ that can strengthen the study at the human population level. For this study, we conducted untargeted metabolomics profile analyses in plasma to identify endogenous and exogenous metabolites associated with clinical AD, and to identify differences by racial/ethnic group and *APOE* genotype.

## Methods

### Participants

The Washington Heights, Inwood Columbia Aging Project (WHICAP) has recruited a representative community based group of individuals aged 65 years and older through a collaborative effort with the Centers for Medicare and Medicaid and through the use of marketing rolls. The composition of the cohort reflected the community surrounding Columbia University Medical Center and comprises of non-Hispanic whites (24%), African Americans (28%), Caribbean Hispanics (48%) and 67% of the cohort are women. After obtaining informed consent, participants are interviewed in either English or Spanish. During each assessment, participants received a standardized neuropsychological test battery and medical interview. Blood was drawn, bar-coded, and brought to the laboratory within two hours of collection for DNA extraction and storage of plasma and serum. All of the medical, neurological, psychiatric, and neuropsychological data collected were reviewed at a consensus conference by clinicians who are experienced in the diagnosis of AD and related dementias. Diagnosis of clinical AD was based on accepted criteria^2^.

For this investigation, fresh plasma was obtained, processed and frozen within two hours from individuals who had been evaluated at least twice to ensure stability of the clinical diagnosis AD and the lack of dementia in healthy controls. These individuals were equally divided between the three ethnic groups: African American, Caribbean Hispanic and non-Hispanic white and between cases and controls (Table 1).

**Table 1.**
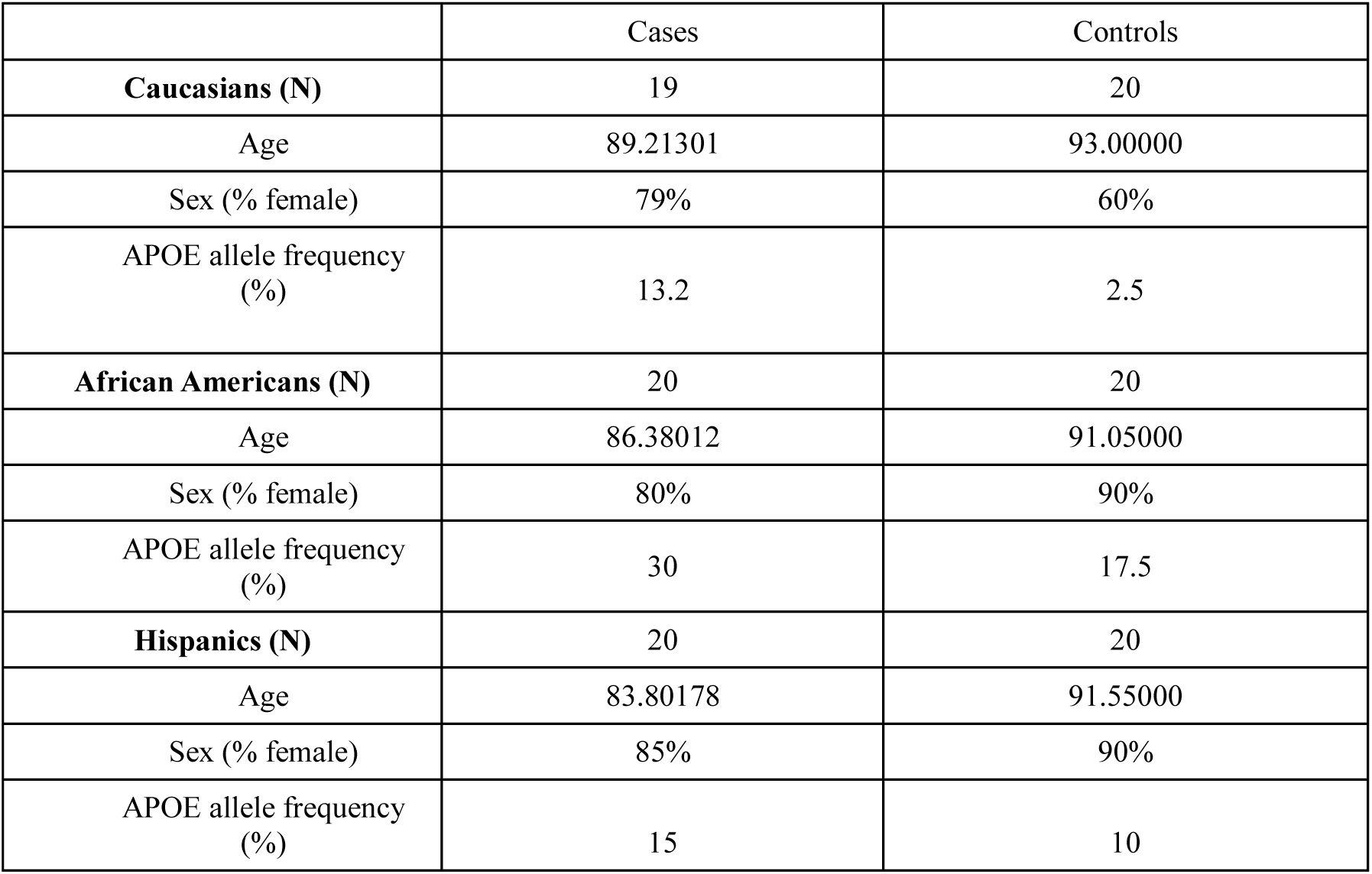
Demographic characteristics of cases and controls analyzed in the study.

|  | Cases | Controls |
| --- | --- | --- |
| **Caucasians (N)** | 19 | 20 |
| Age | 89.21301 | 93.00000 |
| Sex (% female) | 79% | 60% |
| APOE allele frequency (%) | 13.2 | 2.5 |
| **African Americans (N)** | 20 | 20 |
| Age | 86.38012 | 91.05000 |
| Sex (% female) | 80% | 90% |
| APOE allele frequency (%) | 30 | 17.5 |
| **Hispanics (N)** | 20 | 20 |
| Age | 83.80178 | 91.55000 |
| Sex (% female) | 85% | 90% |
| APOE allele frequency (%) | 15 | 10 |

### Metabolomics acquisition and analyses

We acquired metabolites by conducting untargeted metabolomics from the plasma in 119 cases and controls from the WHICAP cohort using previously standardized method^3^. Untargeted LC-based ultra-high resolution mass spectrometry (LC-UHRMS) allowed deep phenotyping of the human metabolome, providing measures of metabolites in most <u>Kyoto Encyclopedia of Genes and Genomes</u> (KEGG) human metabolic pathways^4^. After chromatographic separation using methods previously described^5^, each sample was injected on each column (HILIC with positive electrospray ionization (ESI) and C18 with negative ESI) three times, to obtain three technical replicates per column. Data were processed through a computational pipeline that leverages open source feature detection and peak alignment software, apLCMS^6^ and xMSanalyzer^7^, to create a feature table with the mass-to-charge (*m/z*) ratio, retention time, and the abundance of each ion in each sample^6-8^. The feature table was used for statistical analyses followed by untargeted pathway analysis using the software Mummichog^8^. The LC-UHRMS platform detects over 1,500 chemical signals that arise from core nutrient metabolism, lipids, the microbiome, diet-derived chemicals, pharmaceuticals and environmental contaminants, as well as over 100,000 untargeted features^9^. For simplicity, we refer to the mass spectral features as metabolites. Results described as annotations refer to level 4 and 5 confidence of feature identification by criteria of Schymanski et al^10^. Results described as identifications refer to level 1 confidence (accurate mass, retention time and MS/MS fragmentation matching authentic standards).

### Metabolomics data analysis

Raw metabolite intensities were corrected for batch effects using ComBat^11^. Metabolites missing in over 30% of the samples were excluded from further analysis. Metabolite values missing or of low quality were imputed to half the value of the lowest level of detection for each metabolite^12^. Intensities were quantile normalized, log-transformed and auto-scaled to normalize distributions^13^.

Principal component analyses (PCA) of normalized values were used to estimate stratification by ethnic/racial group, disease status and APOE ε4. Partial Least Squares Discriminant analysis (PLS-DA) implemented in the mixOmics^14^ package was used to conduct supervised dimension reduction analysis to obtain a linear combination of the features that correlated with variability by ethnic/racial group, disease status and APOE ε4 in the metabolomics data.

We tested association of individual metabolites with AD adjusting for age, sex, *APOE*-ε4 and ethnic/racial group. In addition, we tested the association of these metabolites with AD independently in each ethnic/racial group (defined by PCAs and ancestry analysis of GWAS data) to identify both ancestry specific and population-based metabolomics signatures in AD. Then, we tested association of *APOE ε4* with metabolite intensities adjusting for age and sex. For association testing of independent metabolites, we declared nominal significance at P<0.05. Nominally significant metabolites were subsequently tested for pathway enrichment. Pathways enriched for metabolites associated with traits of interest were detected using module-enrichment analysis implemented in mummichog^8^. Statistical significance of enriched pathways were determined using a permutation test adjusting for multiple testing^15,16^.

The project was reviewed and approved on 11/19/2019 by the Columbia University Medical Center, Institutional Review Board (IRB-AAA09804).

## Results

Untargeted metabolomics profiles were completed in 119 individuals that included 40 African Americans, 40 Caribbean Hispanics and 39 non-Hispanic whites from the WHICAP cohort. Using the HILIC column with positive electrospray ionization (ESI), we identified over 9,700 features, of which 5,929 were putatively annotated using xMSannotator. The features did not show batch related variation and a total of 6375 features were present in at least 70% of the samples studied, of which 1,704 metabolites were annotated with xMSannotator confidence level 2 or 3. Using the C18 column with negative ESI, we measured over 6,700 features, of which 3759 were present in at least 70% of the samples.

### Metabolomic profiles of AD

We compared metabolites between AD and controls adjusted for age, sex and ethnic/racial group and found several metabolic features nominally associated with AD (uncorrected p<0.05) measured on both columns (figure 1A, Table S2) as well as unique metabolic profiles (Table 2).

**Table 2.**
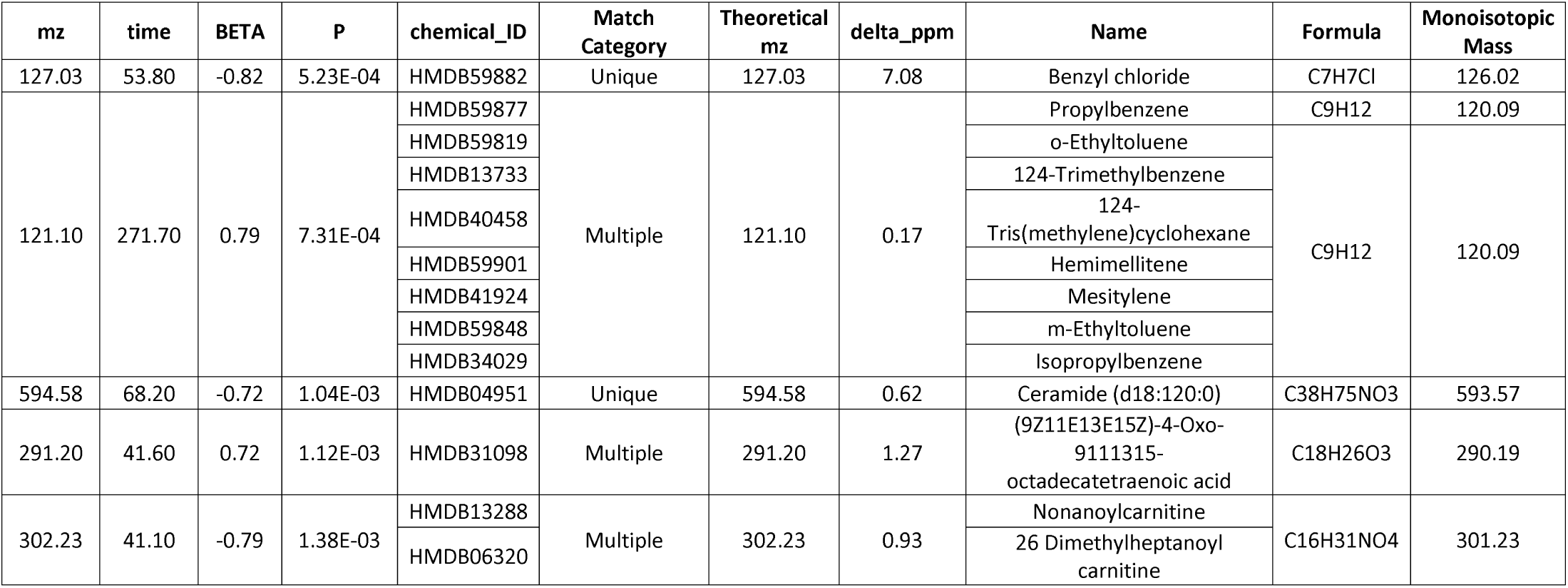
Individual metabolites associated with AD status and annotated with a confidence score>2.

**Figure 1A-D.**
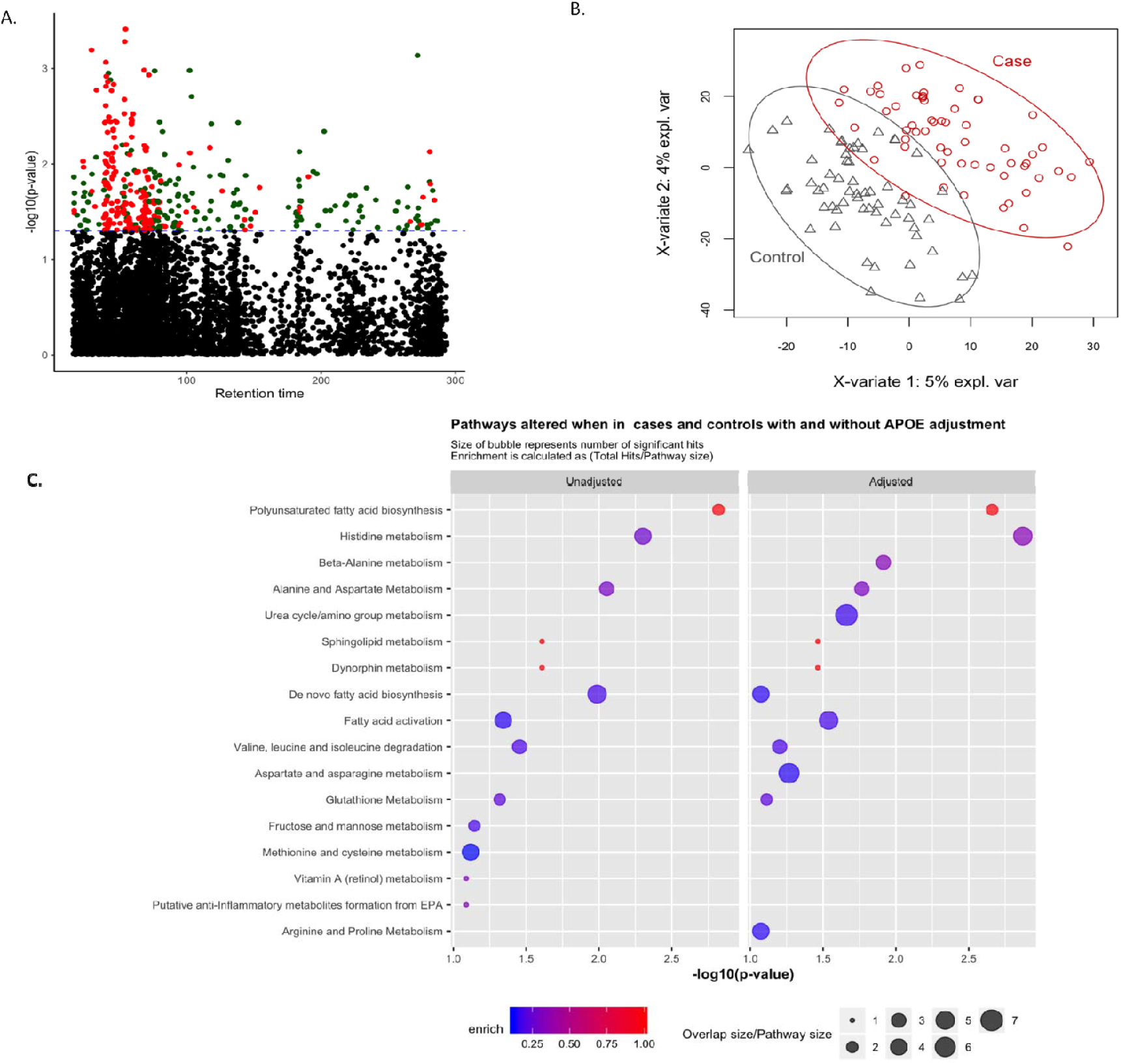
Metabolome wide association study in cases and controls of AD. A. A metabolome wide association study (MWAS) found 382 features altered in cases and controls at nominal significance of p<0.05 In red: features lower in cases, in green features higher in cases: B. A PLS-DA shows metabolomic patterns different between cases and controls (unadjusted for age and sex variables). C. Pathway analysis using mummichog shows most probable metabolic pathways altered between cases and controls.. We tested enrichment of pathways among metabolites that were nominally significant in association with AD (p<0.05) in models unadjusted and adjusted for APOE respectively. Displayed pathways are enriched at p<0.05, corrected for multiple testing.

While none of individual metabolites reached experiment-wide significance after correcting for multiple testing, we identified several metabolites annotated with a confidence level 2 by xmsannotator that were associated with AD at p<10e-03 (Table 2). In particular, we identified benzyl chloride, various adducts of benzenes and toluenes, omega 3 fatty acids, ceramide and carnitines that are associated with Alzheimer’s Disease.

In addition, features from the HILIC positive data indicated that altered polyunsaturated fatty acid biosynthesis, alanine, aspartate, glutamate, glycerophospholipid and sphingolipid metabolism (figure 1C) were also associated with AD metabolome-wide. Data from the C18 negative column implicated altered glycolysis and gluconeogenesis, pyruvate and alanine and aspartate metabolism in cases versus controls of AD. Individual metabolites that contributed to enrichment of these pathways are described in Supplementary Material.

### Metabolomic differences by ethnic/racial group

Within each ethnic/racial group, we found that African Americans with AD had altered glycolysis and amino acid metabolism as well as polyunsaturated fatty acid metabolism. Non-Hispanic whites with AD had altered amino acid, fatty acid, and glycosphingolipid metabolism, and Caribbean Hispanic cases had altered amino acid metabolism (figure 2). Restricting the analysis to controls, we determined underlying metabolic differences that existed in participants based on their ethnic/racial group was not driven by the presence of disease (figure 3A, Table S1).

**Figure 2.**
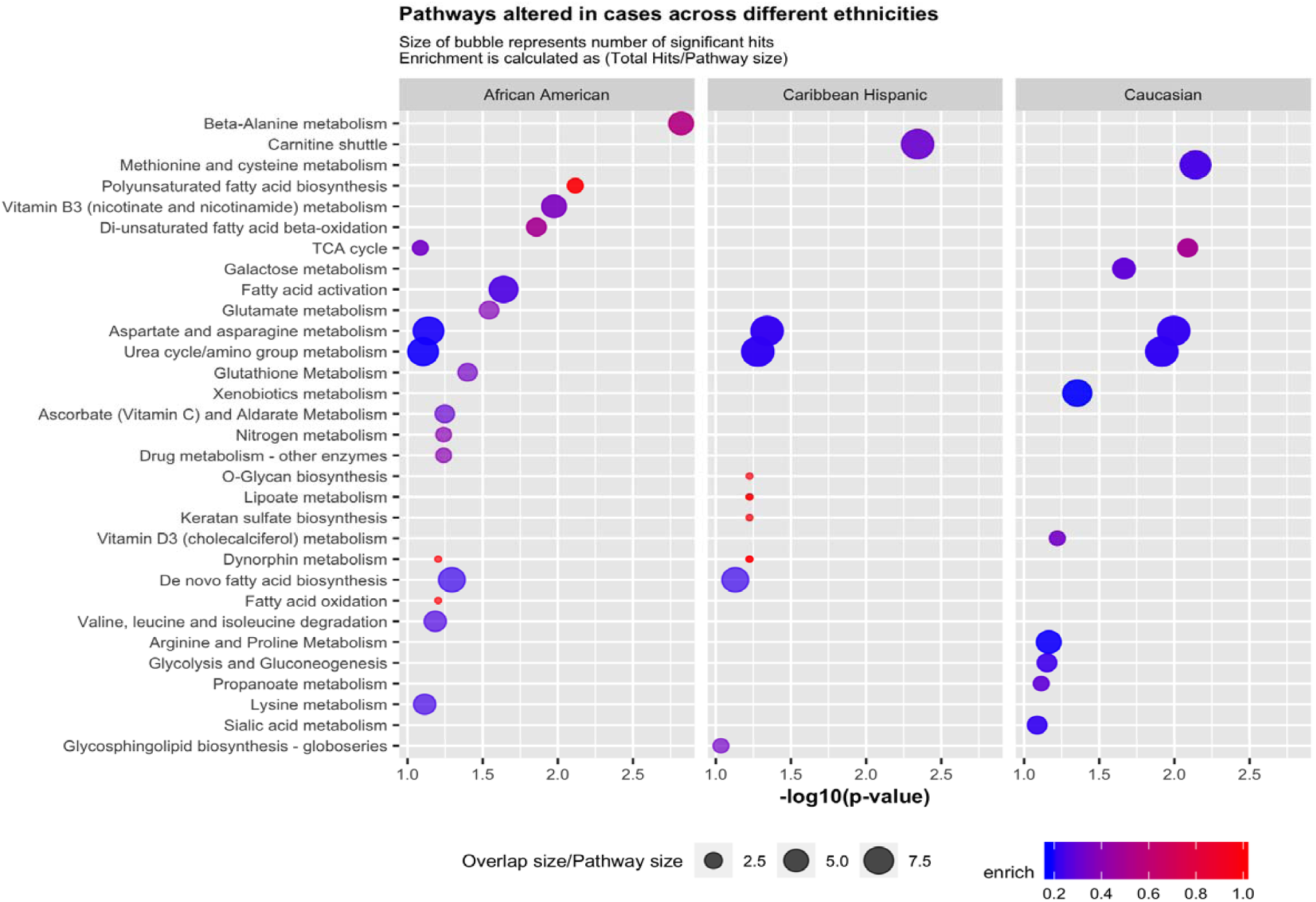
Ethnic differences. A bubble plot that shows pathways altered in controls of the three ethnic groups. Size of bubbles is proportional to overlap size of pathway and p-values are corrected for multiple testing.

**Figure 3.**
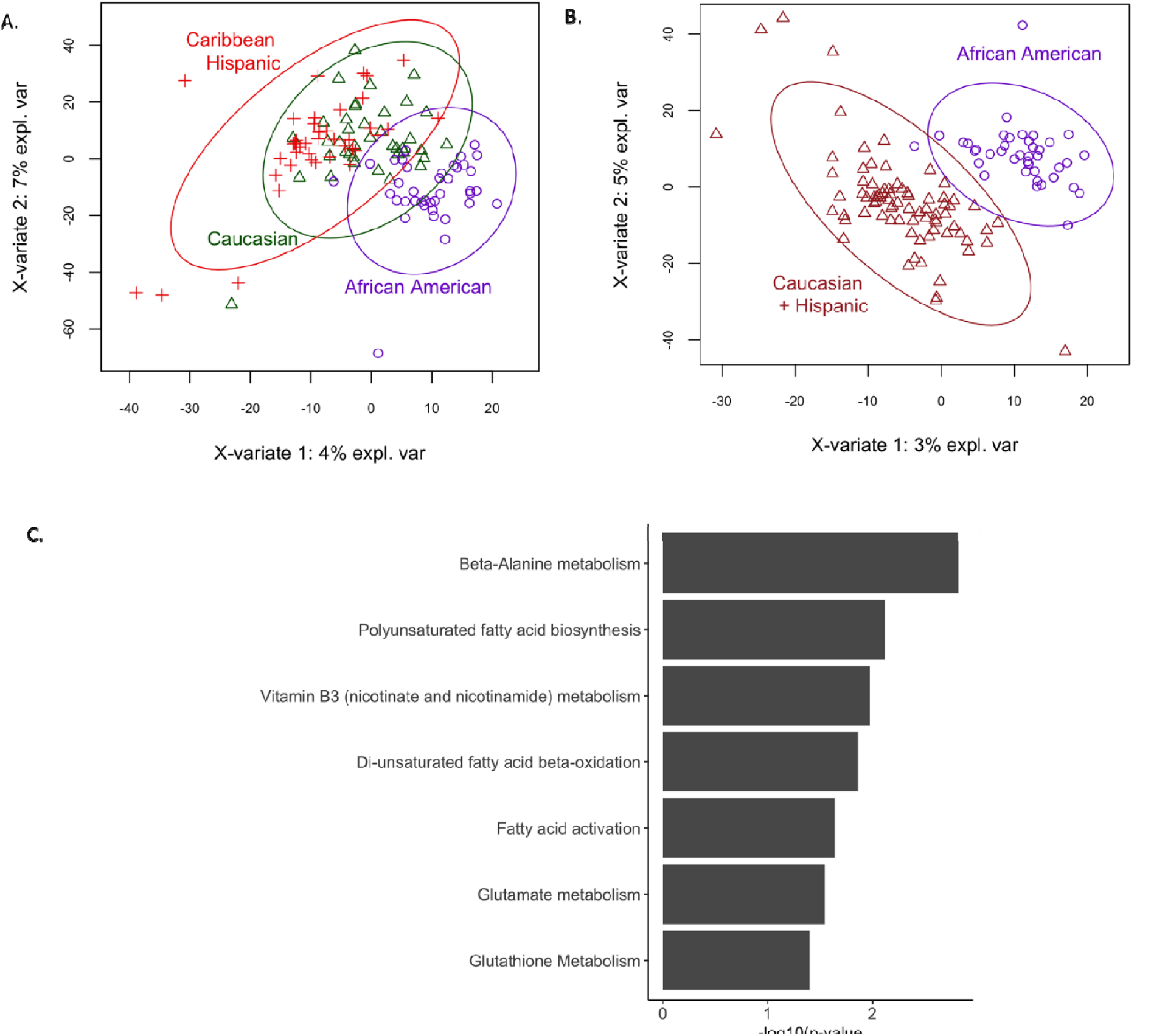
African Americans v others. A. A PLS-DA with all ethnic groups shows a African Americans differ from the Caucasian and Hispanic individuals. B. A PLS-DA the shows differences when comparing AAs with all others. C. Pathways analysis of features that are significantly different in cases and controls of African Americans. D. Distribution of top 3 features significantly different in cases and controls of African Americans in African American and other ethnic groups combined.

Compared with Caribbean Hispanics and non-Hispanic whites, partial least squares discriminant analysis (PLS-DA) suggested features in African Americans clustered differently (Figure 3), including altered amino acid metabolic pathways such as arginine and proline metabolism, asparate and asparagine metabolism, and gylcine, serine, alanine and threonine metabolism. Saturated fatty acid beta oxidation was also identified based on variable importance in the projection (VIP) scores from the PLS-DA (figure 3).

### Metabolomic profiles associated with *APOE* genotype

We also investigated metabolic differences driven by *APOE*-ε4. PLS-DA identified near complete separation between *APOE ε4* carriers and non-carriers (Table S3). In contrast to pathways identified by AD diagnosis, the presence of *APOE*-ε4 was associated with changes in arachidonic acid metabolism, driven by features with putative matches to octadecatrienoic acid and icosatrienoic acid (figure 4A and B), changes in the pentose phosphate pathway, and in chondroitin sulfate. After adjusting for *APOE*-ε4, the pathways associated with AD were largely the same (figure 1C).

**Figure 4.**
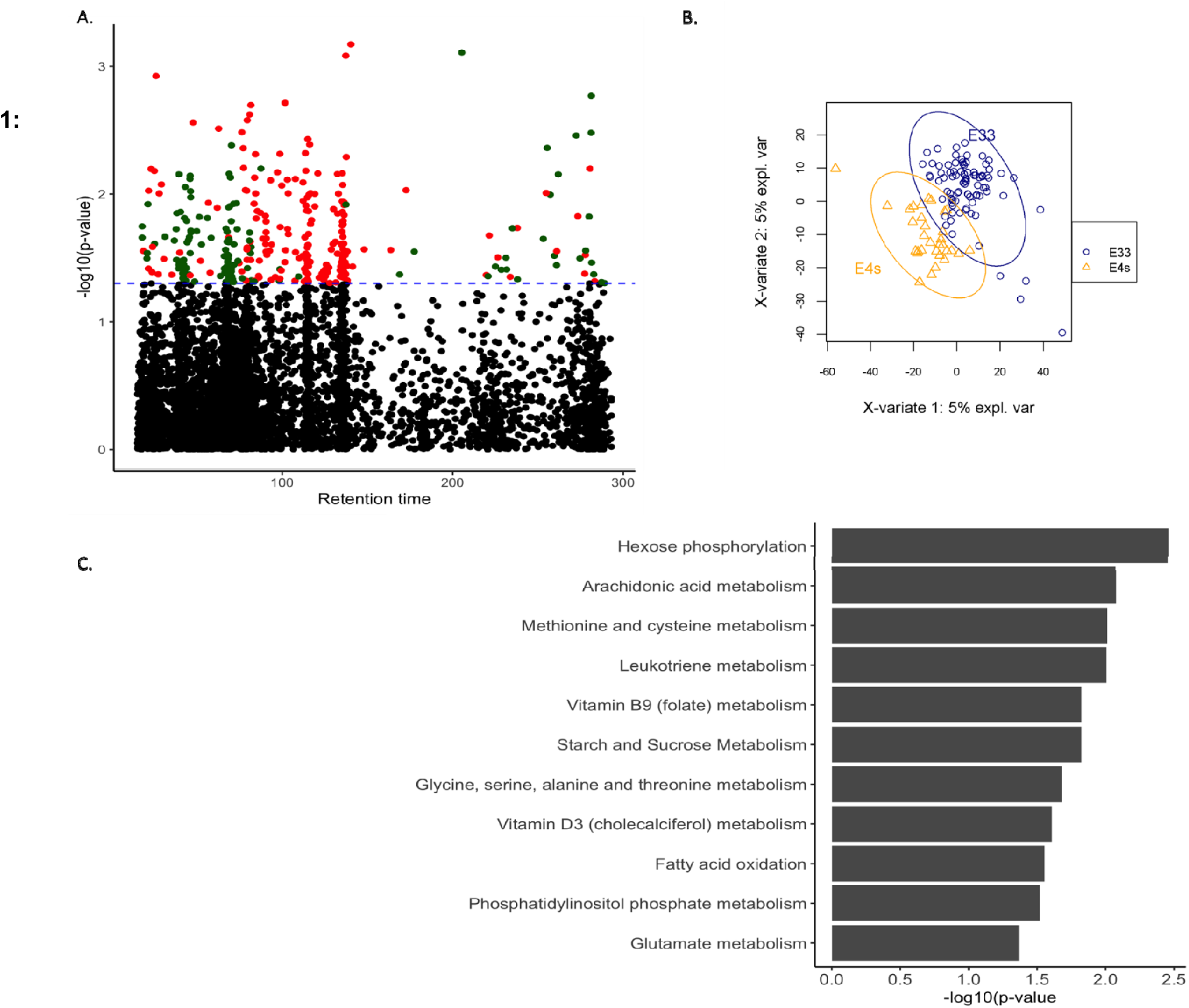
APOE status and metabolic differences. A. MWAS shows features associated with APOE status. B. PLS-DA showing separation between APOE ε4 carriers and non-carriers C. Metabolic pathways enriched in metabolites that are associated with APOE ε4 status at a nominal significance of p<0.05.

## Discussion

We compared untargeted metabolomics in patients with AD and healthy controls, equally divided between African Americans, Caribbean Hispanics and non-Hispanic whites. The goal of this investigation was to identify metabolites associated with disease while adjusting for the strongest genetic risk factor, *APOE* and ethnic/racial group. Therefore, we used a simple case-control design equally stratified by ethnic/racial group.

We found significant differences comparing exogenous and endogenous metabolites in patients with AD and healthy controls. Similar differences were observed in a comparison of metabolic profiling in brain tissue from the middle frontal and inferior temporal gyri in healthy controls, patients with AD or mild cognitive impairment (MCI) in the Baltimore Longitudinal Study^17^. An untargeted approach has also been used to correctly classify patients with mild cognitive impairment or AD when compared with controls with 97.7% accuracy^18^. However, metabolite analyses identified 22 putative biochemical pathways, including interlinked areas of metabolism (polyamine metabolism and L-arginine metabolism).

In this report, we identified several metabolites associated with AD that were altered in the fatty acid biosynthesis pathway. Interestingly, dysregulation of unsaturated fatty acid metabolism was previously reported in brain tissue from patients with AD compared to healthy controls^17^. The disruption in polyunsaturated fatty acid biosynthesis may have several downstream effects: inflammation, oxidative stress, cell maintenance or cell death. Several amino acid metabolism related pathways were also enriched among metabolites that were associated with risk of AD. Lower levels of glutamate and aspartate, and higher levels of glutamine have been observed in the temporal cortex, in postmortem samples from patients with AD^19^. A different cohort also showed elevations in plasma levels of glutamine in AD patients^20^. We identified metabolites involved in dynorphin metabolism associated with AD, consistent with previous studies in postmortem samples from patients^21^. Expression of dynorphin, an endogenous opioid peptide, increases with age and has been associated with cognitive impairment in rodent models^22^.

We found ceramide to be significantly associated with AD, and it has been suggested that ceramide promotes neuronal apoptosis, Aβ accumulation^23^ and has been seen at increased levels in patients with AD and co-morbid conditions^24^. In addition, lower levels of plasma acylcarnitine were found in AD patients compared to controls. Plasma acylcarnitines predict altered fatty acid beta-oxidation, and ketogenesis and have been reported to be depressed in AD patients^25^.Metabolic features also differed significantly in the three ethnic/racial groups, especially among African Americans, despite the individuals being similar in age and residing in the same geographical location. Very few studies have investigated overall metabolic differences by ethnicity/race. Investigators from the Women Health Initiative cohort identified significant differences in metabolite profiles between African Americas and individuals of European ancestry (https://www.ahajournals.org/doi/abs/10.1161/circ.139.suppl_1.MP55). Similar differences in amino acid and lipid metabolism were observed in a metabolic profiling study of serum metabolites in African American and European American patients with bladder cancer^26^. Thus, genetic and metabolic factors could help to explain ethnic/racial and racial disparities in the risk factors associated with AD. Similar pathways emerge when we asses difference by AD or ethnic/racial group suggesting that metabolome profiling from plasma could be used to identify racial differences in disease.

Differences between *APOE ε4* carriers and non-carriers were apparent and remained when analyses were restricted to healthy individuals from all three ethnic/racial groups. Interestingly, metabolites associated with AD were different from those associated with *APOE ε4*. Hexose phosphorylation and arachidonic acid metabolism pathways were most enriched amongst metabolites that were associated with *APOE ε4. APOE ε4* carriers converting to MCI/AD had higher arachidonic acid (AA)/docosahexaenoic acid (DHA) ratios in phospholipids compared to cognitively normal ε4 and non-ε4 carriers^27^. Alterations in plasma arachidonic acid were observed in the brains of ε4 carrier mice compared to non-carriers^27^.

Taken together these results suggest that understanding metabolic heterogeneity in AD pathogenesis may enable identification of biological mechanisms for specific subgroups with the disease. Namely, those carrying an *APOE-ε4* allele and among those of African or Hispanic ancestry. This study demonstrates the ability of untargeted metabolomics to reveal biochemical differences in plasma based on ethnicity/race, the presence of *APOE ε4*, and AD, thus informing study design for optimal power of discovery in larger studies.

## Data Availability

Data will be made available through the National Institute of Aging Genetics of Alzheimer's Disease Storage Site

https://www.niagads.org

## Acknowledgements

WHICAP. Data collection and sharing for this project was supported by the Washington Heights-Inwood Columbia Aging Project (WHICAP, PO1AG07232, R01AG037212, RF1AG054023 and R56AG063908) funded by the National Institute on Aging (NIA) and by the National Center for Advancing Translational Sciences, National Institutes of Health, through Grant Number UL1TR001873. The metabolomics work was supported by U2C ES030163 and R01 ES023839. This manuscript has been reviewed by WHICAP investigators for scientific content and consistency of data interpretation with previous WHICAP Study publications. We acknowledge the WHICAP study participants and the WHICAP research and support staff for their contributions to this study.

## Supplementary Material

**S1.**
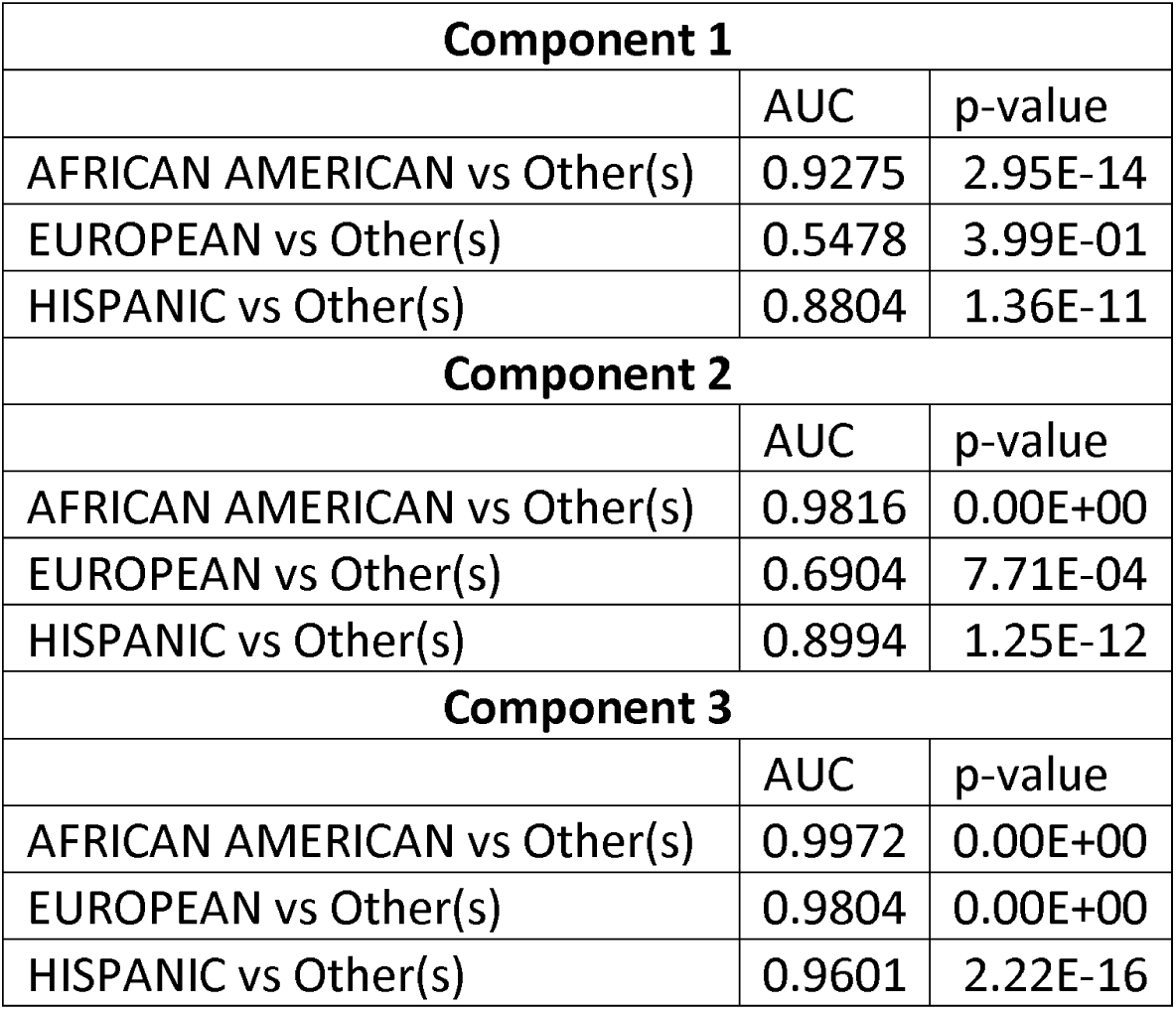
PLS-DA performance in separating three ethnic groups using all 119 individuals (60 AD cases and 59 controls).

| **Component 1** |  |  |
| --- | --- | --- |
|  | AUC | p-value |
| AFRICAN AMERICAN vs Other(s) | 0.9275 | 2.95E-14 |
| EUROPEAN vs Other(s) | 0.5478 | 3.99E-01 |
| HISPANIC vs Other(s) | 0.8804 | 1.36E-11 |
| **Component 2** |  |  |
|  | AUC | p-value |
| AFRICAN AMERICAN vs Other(s) | 0.9816 | 0.00E+00 |
| EUROPEAN vs Other(s) | 0.6904 | 7.71E-04 |
| HISPANIC vs Other(s) | 0.8994 | 1.25E-12 |
| **Component 3** |  |  |
|  | AUC | p-value |
| AFRICAN AMERICAN vs Other(s) | 0.9972 | 0.00E+00 |
| EUROPEAN vs Other(s) | 0.9804 | 0.00E+00 |
| HISPANIC vs Other(s) | 0.9601 | 2.22E-16 |

**S2.** PLS-DA performance in separating 60 AD cases from 59 age-matched healthy controls.

| **Component 1** |  |  |
| --- | --- | --- |
|  | AUC | p-value |
| AD vs HEALTHY | 0.8842 | 4.89E-13 |
| **Component 2** |  |  |
|  | AUC | p-value |
| AD vs HEALTHY | 0.9907 | 0 |
| **Component 3** |  |  |
|  | AUC | p-value |
| AD vs HEALTHY | 1 | 0 |

**S3.** PLS-DA performance in separating APOE4 carriers from non-carriers.

**S3. PLS-DA performance in separating APOE4 carriers from non-carriers**
| **Component 1** |  |  |
| --- | --- | --- |
|  | AUC | p-value |
| E33 vs E4s | 0.9195 | 2.56E-12 |
| **Component 2** |  |  |
|  | AUC | p-value |
| E33 vs E4s | 0.995 | 2.22E-16 |
| **Component 3** |  |  |
|  | AUC | p-value |
| E33 vs E4s | 1 | 0 |

